# Evolution of Life Expectancy and Lifespan Variation in Sub-Saharan Africa

**DOI:** 10.1101/2025.05.20.25328051

**Authors:** Iván Mejía-Guevara, Ursula Gazeley, Doreen Nabukalu, José Manuel Aburto

## Abstract

Despite recent progress, Sub-Saharan Africa (SSA) continues to have the lowest life expectancy and poorest health outcomes globally. While monitoring life expectancy trends is essential, a comprehensive understanding of mortality reduction requires examining age-specific death distributions. Using data from the 2024 Revision of the UN World Population Prospects, we analyze trends in life expectancy and lifespan variation across 49 SSA countries from 1950 to 2023, with projections to 2050, to assess the impact of mortality changes at young, adult, and old ages.

We examine three periods—1950–1979, 1980–2004, and 2005–2023—selected based on key epidemiological milestones, including the emergence of the HIV epidemic and the launch of major global health initiatives around 2000. The impact of mortality reductions varies by age group, resulting in differing patterns of life expectancy gains and lifespan variation. Reductions in child mortality consistently contributed to increased life expectancy and decreased variation, while mid- and old-age mortality showed contrasting trends. Notably, this study offers a novel contribution by examining the impact of HIV-related mortality shocks on lifespan variation.

Our findings challenge conventional frameworks—such as the demographic and epidemiological transitions—in capturing the complexity of mortality change in SSA and call for a reevaluation of prevailing narratives.

## INTRODUCTION

Sub-Saharan Africa (SSA) has the lowest life expectancy and worst health outcomes of any region worldwide. The region’s distinct mortality and health profiles reflect exceptional shifts in demographics and population health over the past 75 years. Persistently high fertility and child mortality rates, the high prevalence of communicable diseases such as HIV, tuberculosis and malaria, and occurrences of conflicts and prolonged social and political instability, have shaped distinct health and mortality landscapes in the region (Defo 2014b; Kagaayi and Serwadda 2016).

Consequently, conventional theoretical frameworks − such as the demographic and epidemiological transitions that have been widely applied to describe and explain changes in population characteristics and disease patterns in Lower-Middle-Income and Upper-Middle-Income Countries may be insufficient to explain complex and evolving dynamics in sub-Saharan Africa (Defo 2014b, 2014a).

Several global targets, including the Millennium and Sustainable Development Goals (MDGs and SDGs), were introduced to improve health and reduce the burden of disease and mortality through coordinated international action. Over the past two decades, some countries have made progress reducing child mortality, HIV prevalence and HIV-related deaths, as well as the burden of other communicable diseases, such as malaria (Rajaratnam et al. 2010; Lozano et al. 2011; UN 2015; Golding et al. 2017; Bennett et al. 2018). However, while significant advances in several countries have been observed, there exists large variation across countries and many failed to meet the MDGs and are off-track to achieve the SDGs by 2030 (IHME 2018; Mejía-Guevara et al. 2019; UN 2000; UN IGME 2025; UNDP 2023; WHO 2025b).

Trends in life expectancy and communicable diseases have been reported before in Africa and worldwide from cross-country comparative perspectives (Anon 2012; Defo 2014b; Masquelier, Reniers, and Pison 2014). Data from the United Nations’ estimates from 1950 to 2010, as well as cause- age-, and sex-specific mortality estimates for 2008 from WHO, involving 57 African countries and used by Defo (2014b), has challenged the suitability of demographic, epidemiological, and health transition frameworks for understanding the African current demographic and health contexts. Unlike Europe and North America, Africa’s mortality, fertility, population growth, and causes of death have followed unique paths in this period (Adetunji and Bos 2006; Baingana and Bos 2006; Bongaarts and Casterline 2013; Defo 2014a; Bongaarts 2017). Those frameworks have been described as insufficient for explaining demographic and epidemiological transformation and predicting future changes in SSA countries due to uncertainties in health, disease, and mortality patterns, and the exceptionalism of the region in those patterns. Hence, their utility to provide a comprehensive representation of the complex situations in Africa for research, planning, and policy purposes has been questioned (Defo 2014b, 2014a).

While life expectancy has been thoroughly studied, a research gap remains in describing long-term longevity patterns along with the variation in the age-at-death distribution in SSA countries. Lifespan variation − variability in length of life − has been described as the ultimate indicator of inequality, as all other measures of disparity are conditional on survival (Tuljapurkar 2010). Lifespan variation is a marker of heterogeneity at the population level that indicates how unevenly distributed mortality improvements are, and at the individual level it is an indicator of how unpredictably age at death is (Aburto et al. 2020a). The significance of lifespan variation is underscored by its potential influence in personal life planning and the structuring of societal measures such as healthcare, insurance, pensions, and related social strategies, which are fundamental for understanding marked differences among various subpopulations (Sasson 2016; Raalte, Sasson, and Martikainen 2018; Nepomuceno et al. 2022). The interplay between life expectancy and lifespan variation has been a focus of study, with historical data pointing to a negative correlation, particularly among medium to high-income nations (Edwards and Tuljapurkar 2005; Smits and Monden 2009; Vaupel, Zhang, and van Raalte 2011; Aburto et al. 2020b). However, recent studies suggest a shift towards positive correlations in certain countries or population groups challenging the overall negative correlation observed in high income countries (Shkolnikov, Andreev, and Begun 2003; Jasilionis et al. 2011; Aburto and van Raalte 2018). Positive associations between changes in life expectancy and lifespan variation have been documented during protracted mortality crises such as those following the dissolution of the Soviet Union (Aburto and van Raalte 2018), increased violence in Latin America (Aburto and Beltrán-Sánchez 2019; García and Aburto 2019), historical epidemics and famines (Vigezzi et al. 2022), and more recently, the COVID-19 pandemic (Aburto et al. 2021).

Given the unique mortality profile of SSA countries, it is a priori unknown how life expectancy and lifespan variation are related in this context. Recent empirical and theoretical efforts aim to elucidate the dynamics between these metrics and ascertain the impact and timing of age-specific mortality on shaping life expectancy and lifespan variation (Aburto and van Raalte 2018; Aburto et al. 2020b; Vigezzi et al. 2022). While life expectancy increases when mortality declines at any age, whether lifespan variation increases or decreases depends on where these improvements occur (Zhang and Vaupel 2009; Gillespie, Trotter, and Tuljapurkar 2014; Aburto et al. 2019, 2022; Martin, Aburto, and Permanyer 2023). This dynamic further complicates the relationship between life expectancy and lifespan variation. Building on this previous literature, this article aims to contribute to the knowledge gap by describing the mortality transition and associated trends in life expectancy and lifespan variation over the last 75 years in SSA. Notably, it provides a novel contribution by examining the impact of mortality shocks from HIV-related deaths on lifespan variation—an aspect that has not been previously studied. In particular, it also shows the contribution of age-specific mortality to changes in those indicators during three defined periods of this mortality transition: 1950-1979, 1980-2004, and 2005-2023.

## METHODS

We used data from 49 SSA countries from 1950 to 2023, and projected data from 2024 to 2050, based on abridged life tables from the 2024 Revision of the United Nations World Population Prospects (WPP) (UN 2024). Life tables were constructed following traditional demographic methods (Preston, Heuveline, and Guillot 2001). Reported age-specific death rates were estimated for each country using mortality data from vital registration systems, censuses, or surveys. The WPP 2024 applied country-specific methods and adjustments depending on the quality and availability of mortality data. For SSA, diverse methods were employed given deficient or lack of data from vital registration systems of countries in the region (AbouZahr et al. 2015). Specifically, child mortality estimates (<5 years old) were obtained from direct methods using full birth histories from survey data—primarily the Demographic and Health Surveys (DHS) and UNICEF’s Multiple Indicator Cluster Surveys (MICS)—or indirect methods with data on children ever born and surviving from available census data or survey data (Hill 2013; UN IGME 2022). As with early neonatal mortality—where data are often not recorded— information on adult mortality is similarly limited and frequently outdated, with some countries lacking data altogether. In this case, estimates were derived using a model-based approach with a Bayesian hierarchical model to approximate the probability of dying between ages 15 and 60. The model incorporated factors such as HIV prevalence, antiretroviral therapy coverage, and child mortality, while also relying on data from neighboring countries for those with limited information. Additional sources included adjusted data on adult household deaths, sibling deaths, and maternal/paternal orphanhood. The specific data sources used for each country are described in the WPP 2024 documentation report (UN 2024).

The estimated age patterns of overall mortality from the WPP 2024 incorporated the demographic effects of HIV-related deaths in numerous countries. This was achieved by integrating estimates derived from HIV/AIDS-specific model life tables, as well as under-5 mortality (ages 0-5) and adult mortality (ages 15-45) figures that were adjusted using UNAIDS data on adult HIV prevalence, including the coverage of antiretroviral therapy (ART) for both children and adults.

### Outcomes

In this paper, we report trajectories of life expectancy at birth and estimate lifespan variation on a country basis. Estimates of life expectancy at birth (denoted as *e*_0_) are directly available from life table data from each country. Life expectancy at birth is the average years a synthetic cohort of newborns is expected to live if individuals from this cohort were to experience the mortality profile observed in a given year during their lives. It is comparable over time and across countries.

We use the standard deviation of the age at death distribution to measure lifespan variation, defined as 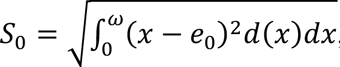, where ω is the highest age group in the population, and *d*(*x*) is the lifetable deaths at age *x*. *S*_0_ has the advantage of being easy to interpret, as it retains the same units as life expectancy values. There exist several indicators of lifespan variation including the Gini coefficient, life table entropy or years of life lost at death (van Raalte and Caswell 2013). They are highly correlated, suggesting that our results would be similar had we used other indicators.

### Analysis of Trends of Life Expectancy and Lifespan Variation

We measure the contribution of each age group to changes in life expectancy and lifespan variation with the linear integral method proposed by Horiuchi, Wilmoth, and Pletcher (2008) (Horiuchi, Wilmoth, and Pletcher 2008), where a change in a continuous function is estimated as:

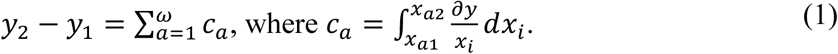

In equation 1, *y*_1_, and *y*_2_ represent the change in the outcome *y* (*e*_0_ or *S*_0_) from times *t*_1_ and *t*_2_; *x_a_*_1_ and *x_a_*_2_ are the mortality rates at age *a* in *t*_1_ and *t*_2_; and *c_a_* is the age-specific contribution to the outcome.

To capture different stages of mortality variation, we consider three periods for analysis in this article: *period 1*, from 1950-79; *period 2*, from 1980-2004; and *period 3*, from 2005-2023. These cut-offs were informed by major epidemiological events or policy goals. The first period ends around the time of the 1978 Alma Ata Declaration, which set the ambitious goal of achieving “Health for All by the Year 2000” (Anon 1978). The second period captures the emergence and escalation of the HIV/AIDS epidemic, first confirmed in Africa around 1980 (Kagaayi and Serwadda 2016), and concludes shortly after the establishment of key global health initiatives. These include the launch of the Millennium Development Goals (MDGs) in 2000— setting as primary targets the reduction in child mortality (MDG 4) and the combat of HIV/AIDS, malaria, and other non-communicable diseases (MDG 6)—, the 2000 UNAIDS World AIDS Campaign, and the establishment of large-scale funding mechanisms such as The Global Fund to Fight AIDS, TB, and Malaria in 2002, and the President’s Emergency Plan for AIDS Relief (PEPFAR) in 2003, which facilitated widespread access to antiretroviral therapy by 2004 (Kebba 2003; Wester et al. 2009; Taylor 2018). The final period (2005-2023) reflects the scale-up of these interventions and the subsequent decline in HIV-related mortality, which peaked in 2005 (HIV/AIDS 2008). The selection of these three periods was also made to characterize countries within the region according to the distinct mortality patterns, the pace of change, and the degree of variation observed throughout each period. The analysis was conducted by gender, given the well-established differences in the age patterns of mortality and survival between males and females (Luy 2003; Colchero et al. 2016).

## RESULTS

### Life Expectancy Trends

Figure 1 shows notable disparities in life expectancy trends across SSA countries. In 1950, most nations exhibited a life expectancy of approximately 40 years. By 1980, only a handful (e.g., Botswana, Cabo Verde, Eswatini, Congo, Gabon, Kenya, among others) had elevated their life expectancy to nearly 60 years. In contrast, some countries (e.g., Angola, Burundi, Chad, Ethiopia, Liberia, among others) remained stagnant around the initial 40-year mark or experienced sluggish growth. In the subsequent period, spanning from 1980 to 2004, many countries faced either stagnation or substantial declines in life expectancy due to conflicts and the emergence of epidemics. Examples include Rwanda in 1994, the First (1997-97) and Second (1998-2003) Congo Wars, and Eritrea in 1998. Notably, from 2005 to 2023, the third period witnessed a resurgence in life expectancy across some countries, with many reaching or surpassing 60 years. However, many of the Southern African countries most affected by HIV (e.g., Eswatini, Lesotho, Namibia, South Africa, Zambia, and Zimbabwe) are beginning to recover, with their trajectories only approaching pre-epidemic levels in recent years. As anticipated, gender differences favor females over males, although the gaps vary markedly between countries and specific years. For instance, in 1998, the life expectancy of males in Eritrea witnessed a pronounced decline due to its conflict with Ethiopia. Another noteworthy instance is the widening gender gap observed in Cabo Verde and Ethiopia during the latest period.

**FIGURE 1.**
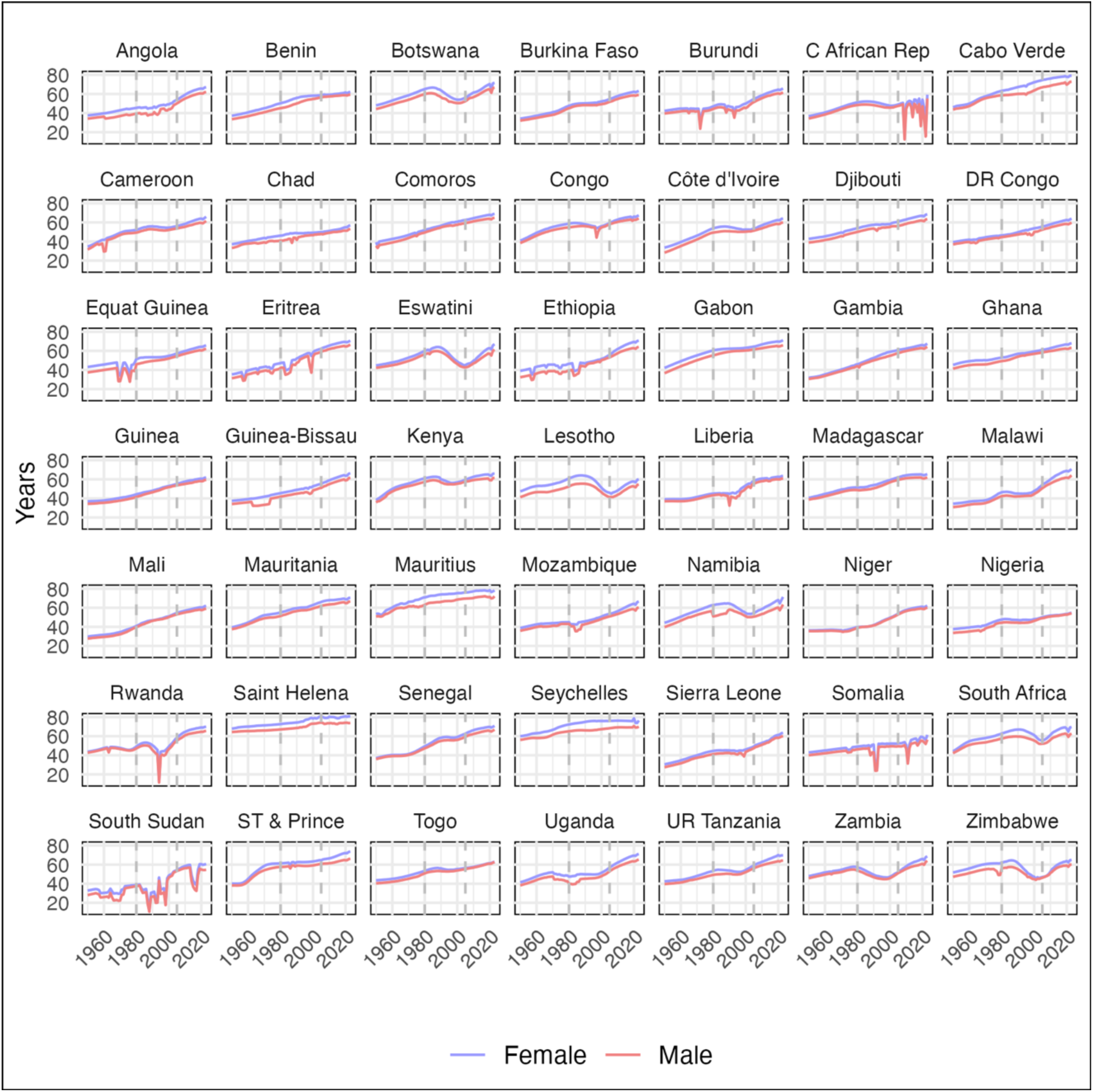
Life Expectancy at Birth (e₀) Trends by Gender in 49 Sub-Saharan African Countries, 1950–2023 **Source:** Authors’ calculations using data from the UN World Population Prospects 2024 Revision.

### Contribution of Age-specific Mortality on Life Expectancy

Figure 2 depicts the contribution of mortality rates from three broad age groups (0-14, 15-59, and 60+) on changes in life expectancy during three different periods. In period 1, 1950-79, panel A shows that the main contributor to life expectancy was the reduction in child and adolescent mortality (age group 0-14), although with variations across countries in terms of magnitude but not significantly across gender groups.

**FIGURE 2.**
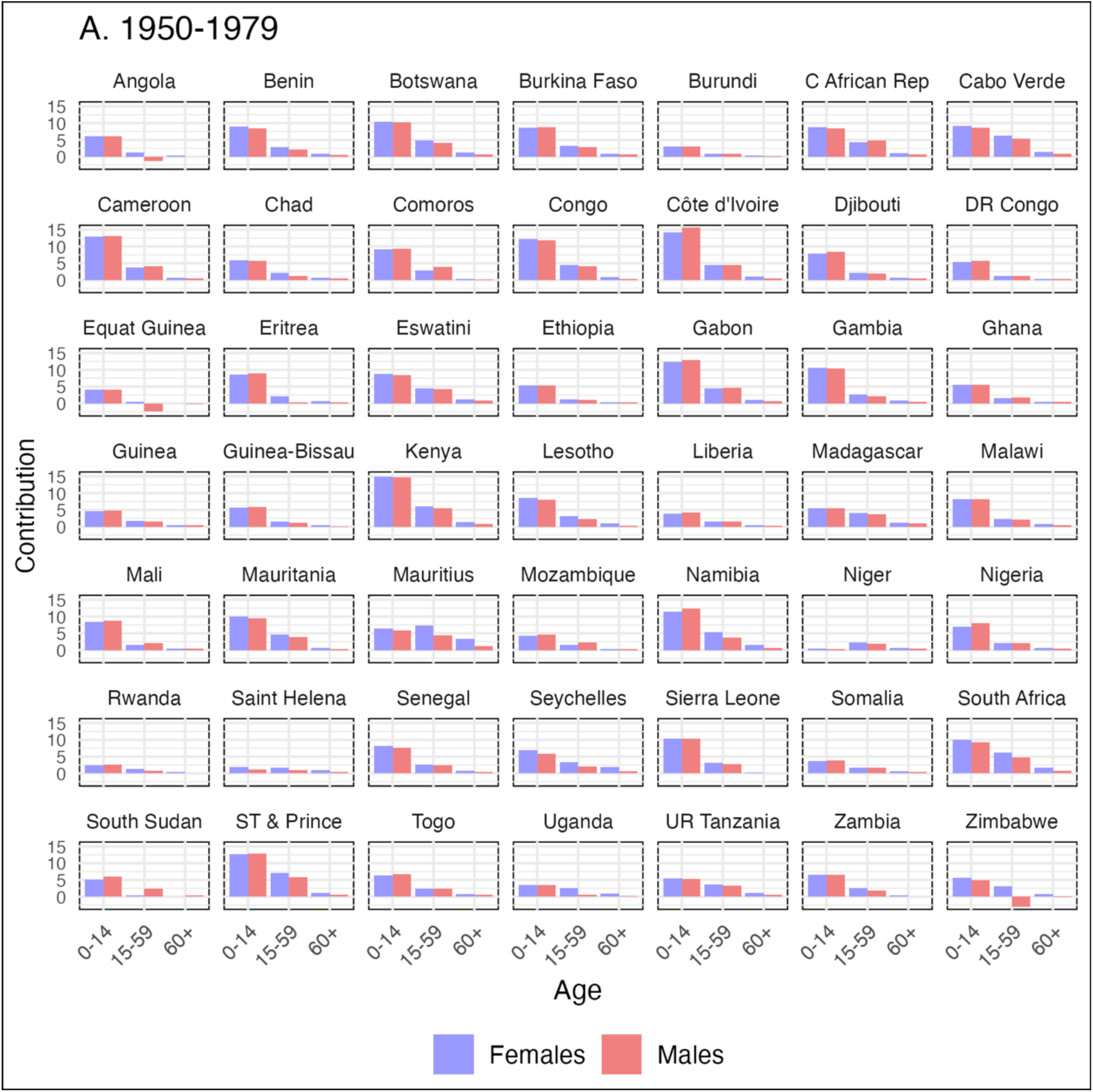
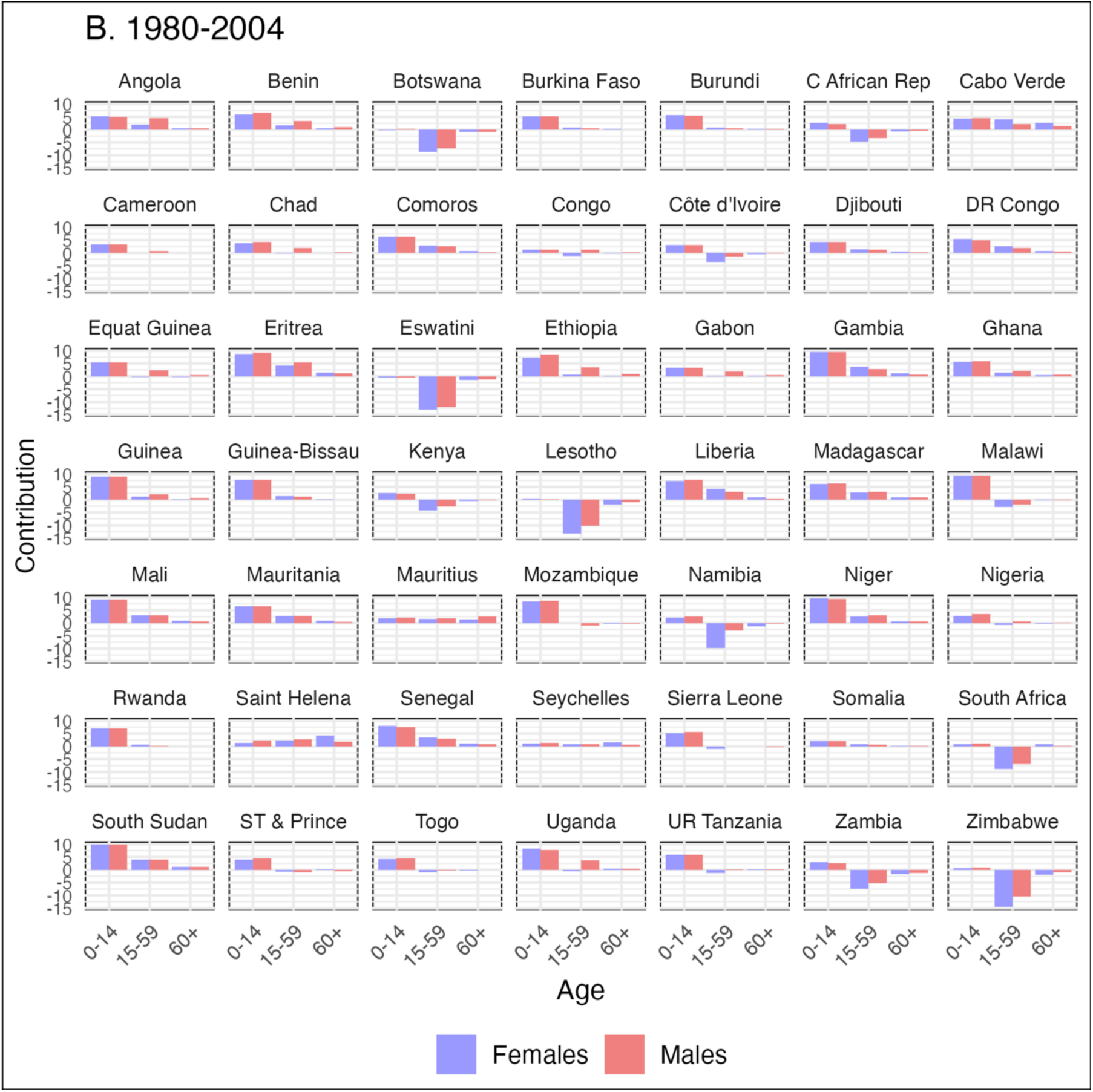
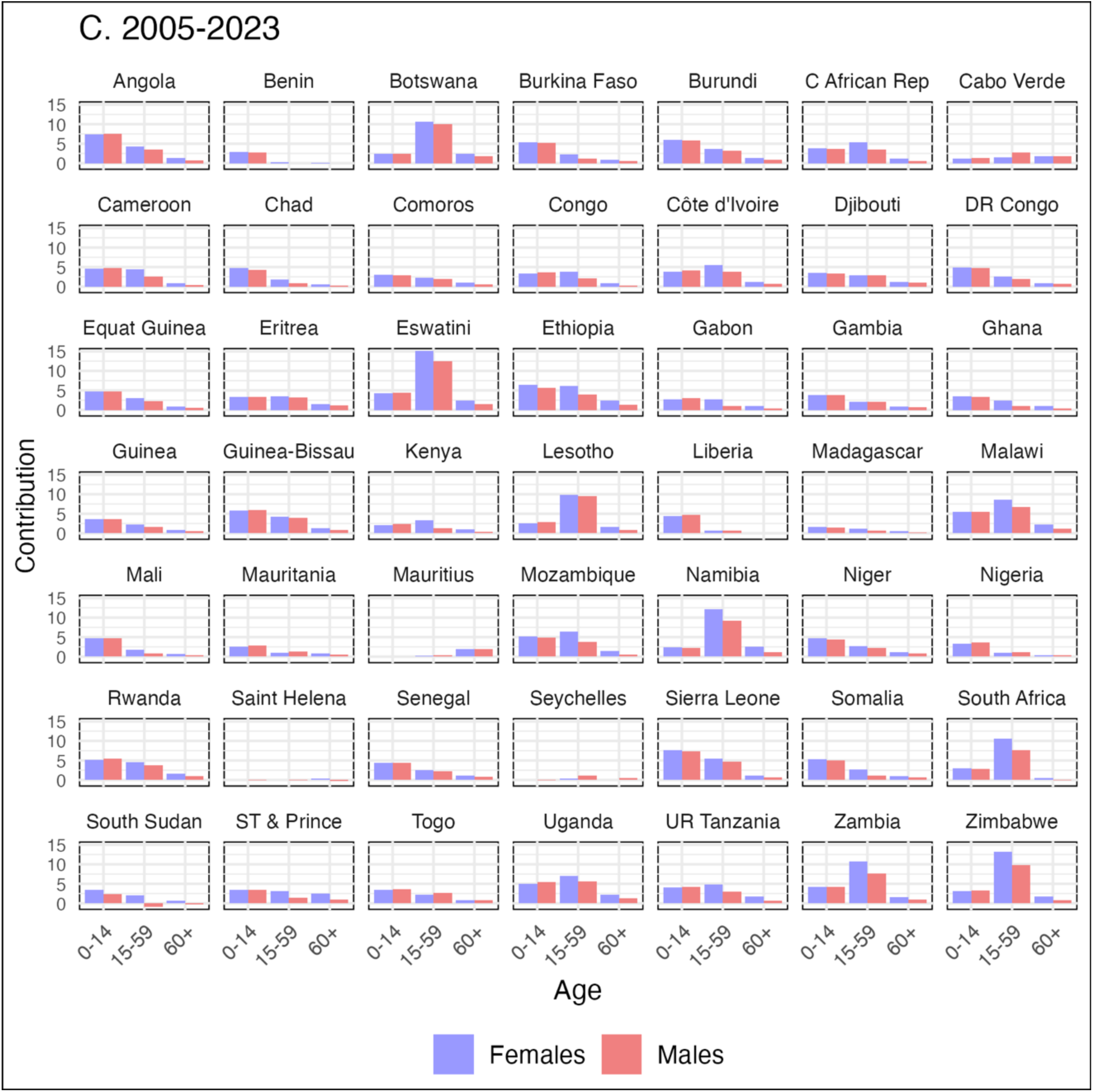
Age-Specific Mortality Decomposition of Life Expectancy at Birth by Time Period, and Gender in 49 Sub-Saharan African Countries Panel A: 1950–1979, Panel B: 1980–2004, Panel C: 2024–2050; Age Groups: 0–14, 15–59, 60 and over **Source:** Authors’ calculations using data from the UN World Population Prospects 2024 Revision.

In period 2, 1980-2004, (panel B), reductions in mortality at ages 0-14 continued but at a slower pace than before while witnessing negative contributions from adult groups aged 15-59 in several countries. This is particularly pronounced in countries severely impacted by the HIV/AIDS epidemic. For example, in Zimbabwe, mortality within this age group led to a staggering 10-year reduction in male life expectancy, and nearly 15-year reduction for females. The impact was substantial in other highly affected countries in Southern Africa like Botswana, Eswatini, Kenya, Lesotho, Malawi, Namibia, South Africa, and Zambia, resulting in approximately 4 to 10 years of lost life expectancy.

Finally, from 2005 to 2023 (panel C), we observed mortality reductions across almost all age groups, leading to increased life expectancy, although in many countries, levels have only now returned to those seen before the HIV epidemic. Notably, the most substantial contributions to these improvements were seen in child and mid-age mortality reductions across most countries, albeit with noticeable variations. To illustrate the case of children, while in Mauritius and Seychelles, the contribution of the 0-14 age group was less noticeable or nearly negligible, this group contributed roughly 7.5 additional years of life expectancy in both Angola and Sierra Leone for males and females.

For mid-age groups, the contribution was particularly notable in nations heavily affected by the HIV/AIDS epidemic, with differences between genders. For instance, we observed a mortality reduction of 15 years for females and 12.5 years in males in Eswatini, 10-11 years in Botswana, while in Namibia, it amounted to 12 years for females and 9 years for males. Similarly, in Zambia, the contribution was 11 years for females and nearly 8 years for males, and in Zimbabwe, it stood at 13 years for females and 10 years for males.

While to a lesser extent, and only in some countries, older groups aged 60 and above also increased their contributions to life expectancy, particularly among females. For instance, old-age groups contributed nearly 3 additional years to female life expectancy and 1 year to male life expectancy in Ethiopia and Sao Tome and Prince.

### Lifespan Variation Trends

Figure 3 illustrates trends in the standard deviation of the age at death (*S*_0_) with large variation between countries. We have identified four general patterns. First, some countries exhibited stagnation at high *S*_0_ levels from 1950 to 2004, followed by a significant decrease afterwards (e.g., Angola, Benin, Burkina Faso, Chad, DR Congo, Ethiopia, Sierra Leone, among others). Second, some countries experienced an initial decline in lifespan variation during 1950-1979, followed by a period of stagnation or increase in the years from 1980 to 2004, and then a return to declining trends from 2005 to 2023 (e.g., Botswana, Congo, Kenya, Lesotho, South Africa, Zimbabwe). Third, certain countries demonstrated a consistent and gradual decrease in lifespan variation throughout the entire period, although there was a deceleration or increases observed in some countries along the way (e.g., Cabo Verde, Comoros, Gabon, Namibia, Togo, Zambia). Fourth, group of countries with sharp reductions in male *S*_0_ resulting from conflicts, including Cameroon civil war 1960, Burundi civil war 1972, Central African Republic conflicts 2012-2020, Liberian First (1989-1997) and Second (1999-2003) civil wars, multiple conflicts in South Sudan, Rwandan genocide 1994, Eritrea & Ethiopia conflict 1998-2000 (seen more in Eritrea’s pattern than Ethiopia), Somalia civil war in 1988, coupled with a severe drought, led to a devastating famine in 1992.

**FIGURE 3.**
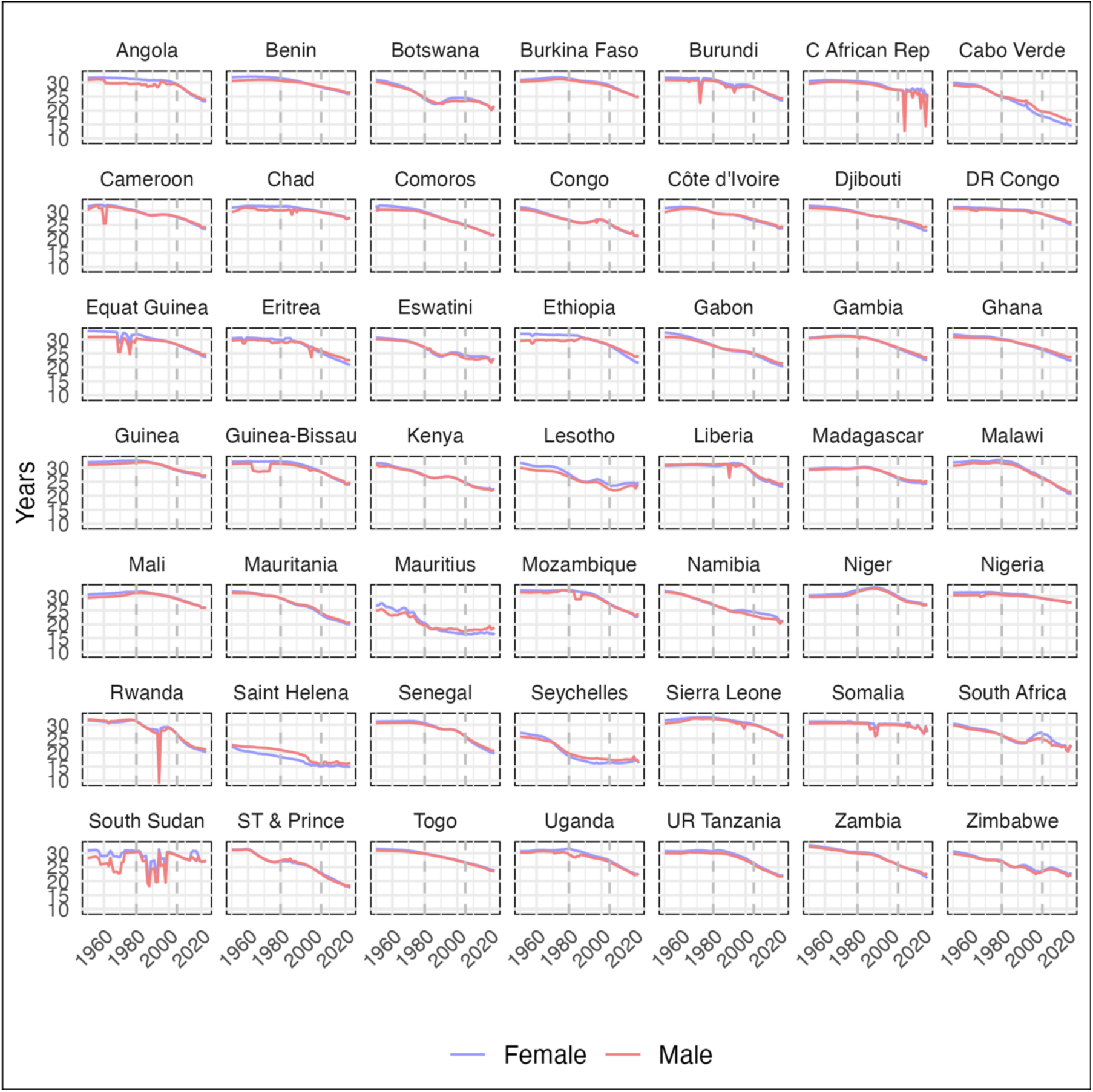
2023Lifespan Variation (*S*_0_) Trends by Gender in 49 Sub-Saharan African Countries, 1950-2023 **Source:** Authors’ calculations using data from the UN World Population Prospects 2024 Revision.

### Age-specific Contributions to Life Expectancy and Lifespan Variation

Figure 4 shows age-specific contributions to life expectancy and lifespan variation by age groups categorized as: Young (0-14), adult (15-59), and old age (60+). We also break down these associations into three distinct time periods described previously. This figure illustrates how general patterns of changes of life expectancy and lifespan variation due to these age groups relate to each other and whether they are consistent with previous findings in the literature. Panel A shows contributions to changes driven by the young age group (0-14). Most countries have experienced sizable gains in life expectancy and reductions in lifespan variation due to improvements in mortality at younger ages. It is evident that there is a negative association between the impact of child and adolescent mortality and the change in life expectancy (*e*_0_) relative to the change in lifespan variation (*S*_0_), regardless of the period. This relationship has become more strongly correlated over time, with the most pronounced effect observed from 2005 to 2023 as youth survival rates improve.

**FIGURE 4.**
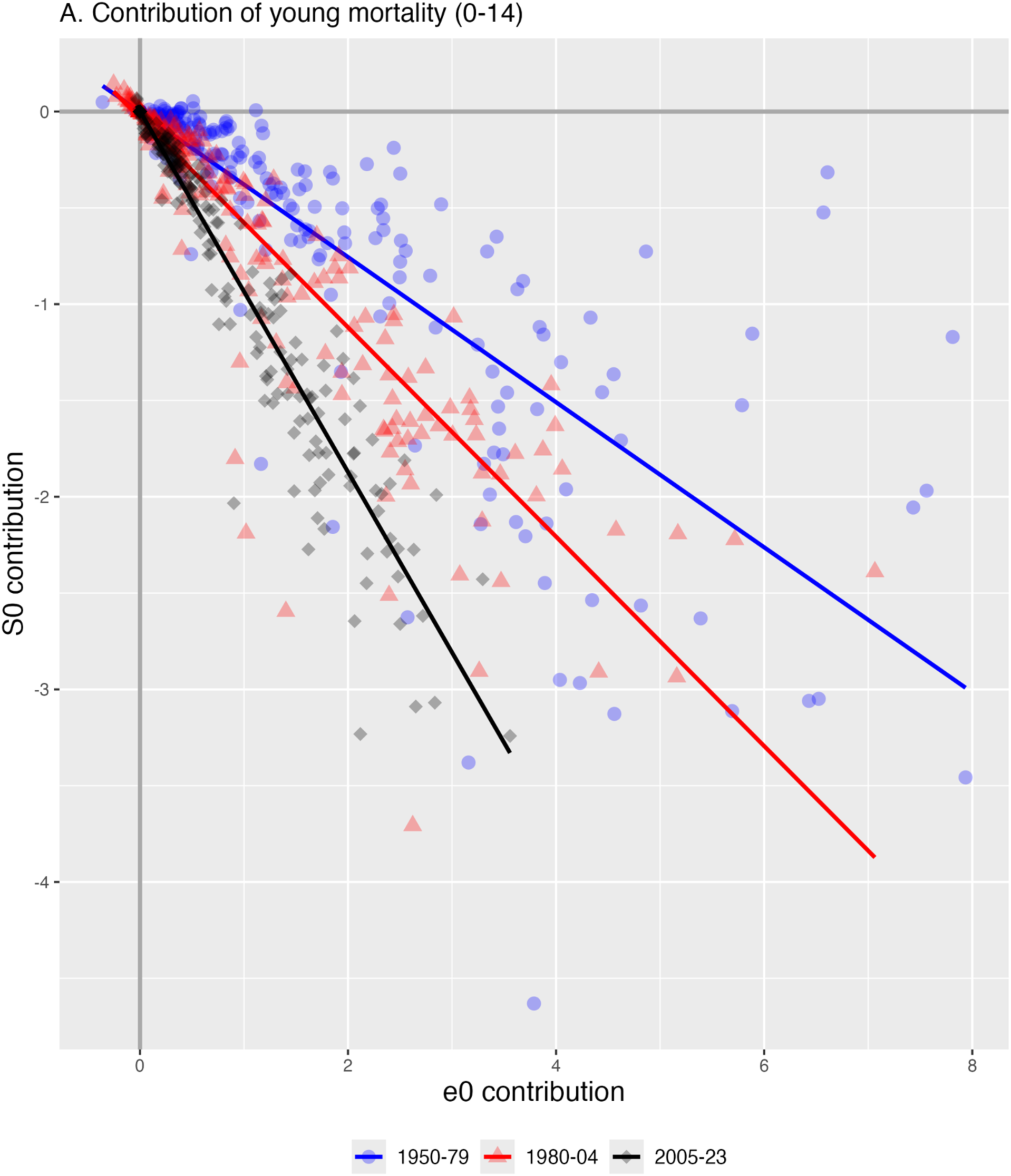
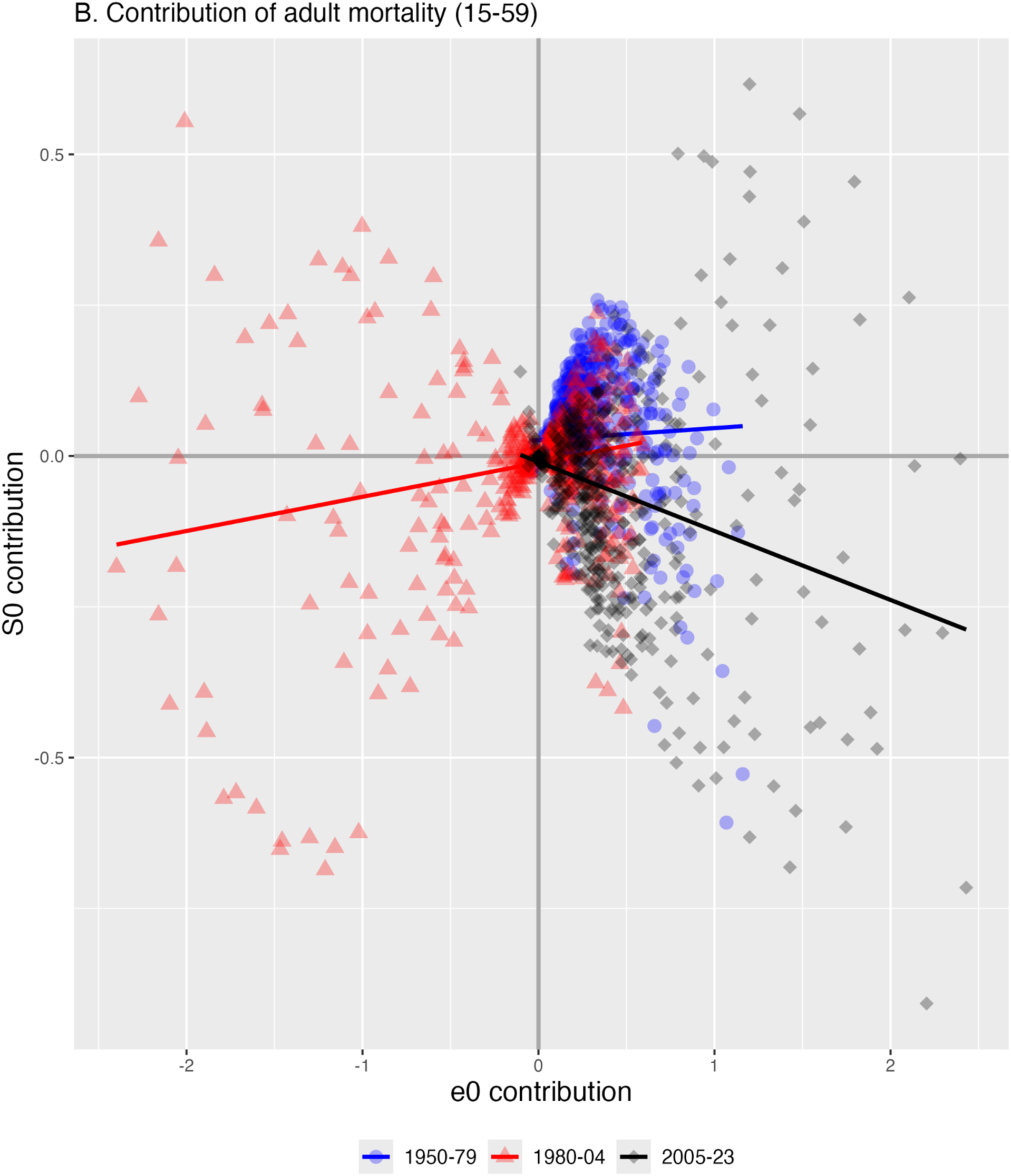
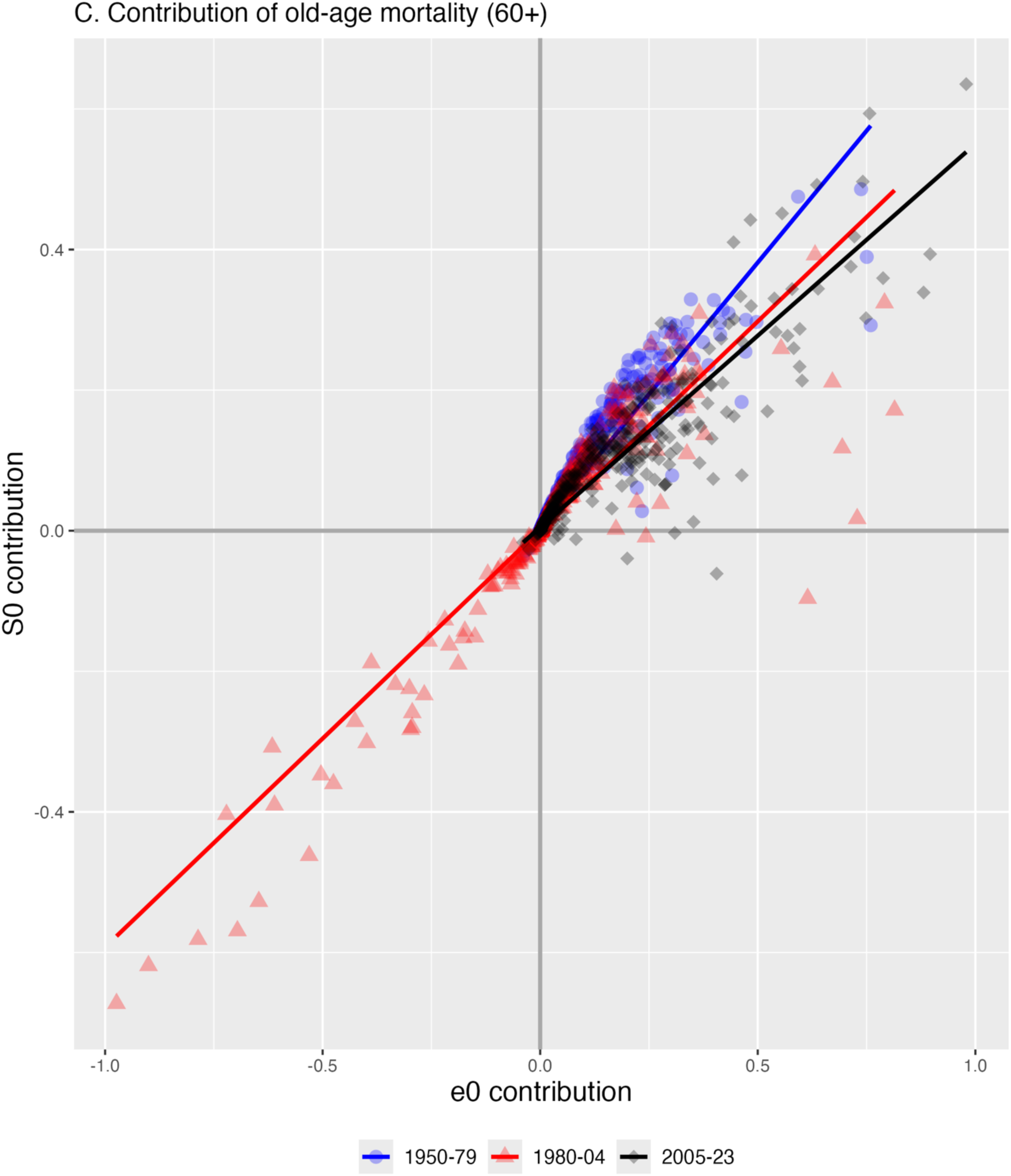
Association Between Age-Specific Mortality Contributions to Life Expectancy (e₀) and Lifespan Variation (S₀) by Time Period in 49 Sub-Saharan African Countries Time Periods: 1950–1979, 1980–2023, 2024–2050 Panel A: Age Group 0–14, Panel B: Age Group 15–59, Panel C: Age Group 60 and over **Source:** Authors’ calculations using data from the UN World Population Prospects 2024 Revision.

Panel B reveals the discordant relationship between *e*_0_ and *S*_0_ at ages between 15 and 59. Normally, an increase in life expectancy coincides with a decrease in lifespan variation. This panel shows that a large proportion of positive changes in life expectancy are followed with increases in lifespan variation (and vice versa) across all periods. This panel also shows that the period from 1980-2004 was marked by substantive losses in life expectancy in this age group. Period 3 resembles what we observed for child and adolescent mortality in Panel A, reflecting a period of sustained improvements in adult mortality in most countries within the region, although some have experienced increases in lifespan variation in parallel.

Panel C depicts a consistent positive relationship between the contribution of old age mortality, regardless of the period. Gains in life expectancy due to older age mortality are usually translated into increases in lifespan variation, while losses in life expectancy translate into decreases in variation.

### Potential Trajectories Through 2050

Figure 5 presents a hypothetical scenario based on projected life tables from 2024 to 2050 from the WPPs, replicating the relationship between the contribution of mortality to *e*_0_ and *S*_0_ across the three different age groups. The figure shows continuous changes in mortality dynamics across age groups. Young mortality still contributes to a negative correlation between *e*_0_ and *S*_0_due to its contribution to increasing *e*_0_ and reducing *S*_0_. However, this relationship varies significantly across countries. For mid-age groups, the progress is notable, with a negative and more homogeneous relationship across all countries. Finally, for older age groups, the relationship remains positive due to their increasing contribution to *e*_0_, but also increasing *S*_0_for these age groups.

**FIGURE 5.**
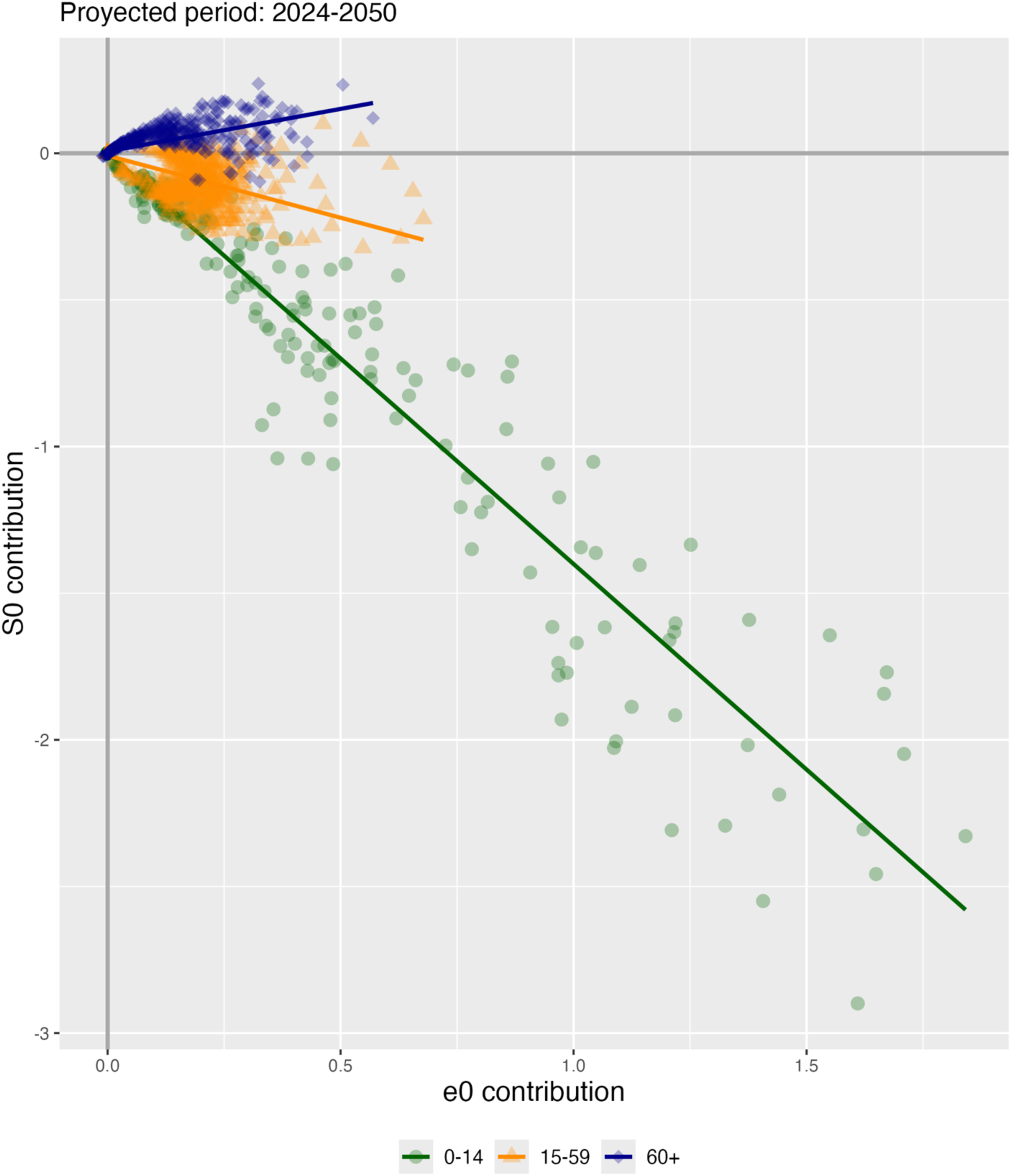
Association Between Expected Age-Specific Mortality Contributions to Changes in Life Expectancy (e₀) and Lifespan Variation (S₀), 2024–2050 in 49 Sub-Saharan African Countries **Source:** Authors’ calculations using data from the UN World Population Prospects 2024 Revision.

## DISCUSSION

This paper analyzes trends in longevity and lifespan variation in 49 SSA countries from 1950-2023. The analysis is stratified by gender and three periods from 1950-79, 1980-2004, and 2005-23, which align with the policy and epidemiological phases outlined earlier, particularly the impact of global health initiatives introduced in the early 2000s.

### Trends in Life Expectancy and Lifespan Variation

In 1950, life expectancy in most SSA countries was around 40 years, and only a few countries saw life expectancy rise to nearly 60 years by 1980. Between 1980 and 2004, life expectancy stagnated or declined due to social and political unrest as well as the HIV epidemic and tuberculosis − the leading cause of death among people with HIV. In the early 1980s, the first cases of AIDS were identified, and the HIV itself was isolated—the identification and extraction of the virus from biological samples— in most SSA countries (Serwadda et al. 1985; Kagaayi and Serwadda 2016). From 2005 onwards, there was a general increase in life expectancy, with many countries surpassing 60 years, although others only returned to pre-HIV epidemic levels. Gender disparities favored females over males during all periods but with variations across countries and times. For instance, between 1980-2004, life expectancy losses were substantially larger for women than men in countries with a high prevalence of HIV/AIDS, such as Botswana, Eswatini, Lesotho, Namibia, South Africa, Zambia, and Zimbabwe. This pattern aligns with the high HIV incidence among adolescent girls and young women previously reported (Birdthistle et al. 2019; UNAIDS 2014). In contrast, in countries not as significantly impacted by the HIV epidemic countries, life expectancy losses were negligible, or no sex differences were observed, throughout the entire period, such as Mali, Rwanda, Seychelles, Nigeria, São Tomé and Príncipe.

Trends in the standard deviation of age at death also varied, with some countries showing high variation until 2004 followed by a decrease, others fluctuating until 2004 and then declining, a third group consistently reducing lifespan variation over the whole period, despite some disruptions/increases, and a fourth group showing significant variation among males in countries affected by conflicts or civil unrest. Apart from these gender differences observed in the fourth group, no further gender differences were observed in most countries, except for a few cases where lifespan variation is consistently lower for women than for men throughout the entire period (e.g., Cape Verde, Saint Helena). Excluding conflicts, our findings of minimal gender differences in lifespan variation in most countries contrast with results from other regions and more general contexts, where females typically have longer and less variable lifespans than males (Colchero et al. 2016).

### Age Decomposition of Life Expectancy and Lifespan Variation Over Time

During the initial period (1950-1979), improvements in child survival was the main contributor to the increase of life expectancy and reduced variation. For instance, in Botswana and Kenya, young survival contributed 10 and 15 years of life expectancy (Figure 2, panel A), and 6.6 and 5.9 in reduced variation, respectively (Figure S2, panel A). The WHO and UNICEF Expanded Programme on Immunization launched in 1974, initially focused on protection against six childhood vaccine-preventable diseases (Bacillus Calmette-Guerin (BCG), diphtheria, pertussis, tetanus, polio and measles), likely contributed to these improvements (WHO 2025a). However, in some countries, midlife survival also contributed to life expectancy gains - for example in Namibia and South Africa, particularly among women with 5.3 and 6.2 years (Figure 2, panel B), respectively, despite much lower contributions to reduced lifespan variation (Figure S2, panel B). This can be a result of improvements in mortality in an age group where mortality decrease is translated into both increases and decreases in lifespan variation, offsetting each other. There exists a ‘threshold age’ below which reductions in mortality contribute to reductions in lifespan variation, while improvements above this age increase variation in lifespans. We hypothesize, that for the case of SSA countries, this age falls at younger age groups that it usually does in high income settings.

Further improvements in child survival occurred in the second period, from 1980 to 2004. The widespread promotion of oral rehydration therapy (ORT) to reduce child diarrheal deaths (Victora et al. 2000), along with advancements in malaria control – such as the introduction of artemisinin-based combination therapies (ACTs) in the 1990s and strengthened global coordination through the Roll Back Malaria partnership launched in 1998 (Thellier, Gemegah, and Tantaoui 2024) – may have contributed to the further reduction in child mortality during this period (Thellier et al. 2024). However, 1980-2004 was also a period of stagnation or increased mortality in middle-aged groups, which contributed to explaining disruptions in overall survival and increased variation in many countries. The impact was substantial in countries heavily affected by the HIV/AIDS epidemic, where declines between 4 to 14 years of life expectancy were observed in the most affected countries, notably Botswana, Eswatini, Lesotho, Namibia, Zambia, and Zimbabwe. This period has been pivotal in shaping the epidemiological and demographic landscape of Africa.

During the last period, 2005-2023, the decline in mortality renewed or accelerated in many countries, particularly with the scale-up of effective malaria control interventions (WHO 2021; Yé et al. 2017), including the widespread distribution of insecticide treated nets (Thellier et al. 2024), expanded immunization programs (Bobo et al. 2022), and better access to antiretroviral treatment for HIV/AIDS (Kharsany and Karim 2016). Programs like the Global Fund to Fight AIDS, Tuberculosis, and Malaria and the President’s Emergency Plan For AIDS Relief (PEPFAR) have played crucial roles in health interventions in the region (Bendavid et al. 2012). These have funded programs such as SAFE (Surgical Circumcision for AIDS Free Era), Prevention of Mother to Child transmission (PMTCT), and DREAMS, successful interventions in combating the HIV/AIDS epidemic in the region (Saul et al. 2018; UNAIDS 2014). Notably, five exemplar countries—Kenya, Rwanda, Tanzania, Senegal, and Uganda—, that are on track to achieve the under-5 Sustainable Development goal targets (Mejía-Guevara et al. 2019), demonstrated how child mortality reductions have been central to life expectancy gains, with the 0–14 age group contributing between two and six years to overall gains (You et al. 2015; Masquelier et al. 2018). Their success reflect diverse strategies, from improved healthcare access, insurance, performance-based financing, and effective use of foreign investment in Rwanda to improve data systems, public health infrastructure, community ownership, and donor coordination in Senegal (Amoroso et al. 2018; Hirschhorn et al. 2025). Meanwhile, countries heavily affected by the HIV/AIDS epidemic, such as Malawi, Botswana, and Eswatini, have made progress in life expectancy for those aged 15-59—ranging from 6 to 15 years—, although recovery was not uniform across all nations and most have only recovered pre-HIV pandemic levels.

These results suggest that in SSA mortality in younger and middle-aged populations has a more significant impact on life expectancy than mortality in older age groups. This reflects the region’s protracted demographic transition, characterized by persistently high fertility and child mortality (United Nations 2022). As a result, SSA is expected to experience an extensive period of demographic dividend—a window of opportunity driven by a growing share of the working-age population—although with variation in the pace of demographic transition among SSA countries (Bloom, Kuhn, and Prettner 2017; Bongaarts 2017; Mason et al. 2017; UN IGME 2022).

### Shifts in the Age-Specific Correlation Between Life Expectancy and Lifespan Variation

An important finding of this study indicates that while most of the mortality variation has occurred at younger ages, there is a disruption in the association at older ages, but particularly in mid-ages where no correlation between improvements in life expectancy and lifespan variation has been consistently identified across countries. The first point aligns with those of Aburto et al. (2020), who demonstrated that improving survival in the youngest age groups strengthens the relationship between life expectancy and lifespan variation. Our analysis reveals a robust negative correlation between the impact of child and adolescent mortality on these two indicators, particularly pronounced in recent years because of the remarkable improvements in child and adolescent survival in the region. Regarding older groups, the lack of correlation between life expectancy and variation in mid-age mortality has been well-documented in the literature, especially in cases of protracted crises such as the dissolution of the Soviet Union (Aburto and van Raalte 2018; Nigri, Barbi, and Levantesi 2021), a period when several factors seem to be at play simultaneously, including economic and social instability, high rates of tobacco and alcohol consumption, poor nutrition, depression, and the deterioration of the healthcare system (Notzon et al. 1998). In addition, political instability, continuous violence, and wars in several countries have significantly impacted mid-age mortality rates. Recent literature indicates a strong association between violence and lifetime uncertainty, showing that violence not only shortens lives but also makes length of life less predictable (Aburto et al. 2023). In the case of SSA, a surge in midlife mortality was notable in the final two decades of the twentieth century. However, improvements in old-age mortality are not as apparent in our SSA data. The decrease in variation and associated reversal in the coefficient of variation of age at death in countries that experienced violent conflicts align with Vigezzi et al.’s findings on historical crises (Figure S1 in the Supplementary Material) (Vigezzi et al. 2022). This is likely due to the intense mortality shock affecting all age groups, causing the distribution to shift to the left. Similar patterns have been observed in other studies, where recent violent conflicts and high homicide rates in Latin America coexist with improvements in other causes of death (Aburto and Beltrán-Sánchez 2019; García and Aburto 2019).

Although there is uncertainty about how these trajectories will evolve in the coming years, our analysis using projected life tables for the region indicates a convergence in the relationship between the mean and variation of age at death in most countries. Specifically, the results suggest that child mortality will continue to contribute to the inverse association of deaths by 2050, with significant differences across countries. This aligns with previous findings that only a few countries will meet the SDGs by 2030, and many will not achieve them even by 2050 (Mejía-Guevara et al. 2019). Notably, the patterns and contributions of mid-age mortality are expected to change significantly, resulting in a negative relationship between life expectancy and age at death in almost all countries. As the aging transition progresses slowly, this relationship remains positive for older age groups but to a lesser extent than in previous periods. This is due to the conflicting dynamics of larger contributions to life expectancy along with the increasing lifespan variation for these age groups.

### Mortality Patterns in sub-Saharan Africa Challenge the Relevance of Transition Frameworks

Taken together, our findings challenge the adequacy of conventional transition frameworks, such as the demographic and epidemiological transition theories, to describe and explain the dynamics of mortality change in sub-Saharan Africa from 1950 to 2023. The distinct and highly heterogeneous trends in life expectancy, causes of death, and lifespan variation, instead are suggestive of exceptionalism in the region’s mortality trajectories. The demographic transition theory, for instance, assumes a long-run, irreversible mortality decline, and fails to anticipate the substantial reversals observed in many countries in SSA, particularly during the 1980-2004 period as a result of the HIV epidemic (Defo 2014b). Similarly, the deterministic, sequential stages of the epidemiological transition as originally proposed (Omran 2005), do not account for overlapping transitions in causes of death or the dual burden of infectious and non-communicable diseases. Nor does the predicted displacement of infectious disease by degenerative diseases account for decline in HIV-related mortality alongside the protracted high HIV prevalence from 2005-2023. Finally, both demographic and epidemiological transition theories assume increasing homogeneity in mortality patterns over time. However, contrary to these predictions of convergence and compression in the age at death distribution (Robine 2001), our analysis reveals diverse and often diverging trends in lifespan variation over time and between countries. This is reflected in the heterogeneous trajectories of the mean and variation in the age at death, and in the evolving relationships between these dimensions as survival improves at younger and midlife ages in recent years. For instance, in some countries, like South Africa, the rise of non-communicable diseases alongside persistent communicable diseases contributes to the increasing mortality in the elderly, indicating a complex health transition phase requiring dual health system preparedness (Modjadji 2021; Wong et al. 2021). Overall, these patterns underscore the delayed and disrupted nature of the mortality transition in many sub-Saharan African settings (Defo 2014b). Ultimately, our findings suggest that the utility of conventional transition theories to meaningfully infer current and future population characteristics in SSA is limited (Caldwell 1998; Defo 2014b; Sudharsanan et al. 2022), and that considerable uncertainty surrounds the region’s future demographic and health trajectories.

### Limitations

This work has some limitations to consider. First, the life table data from the WPP for SSA countries are derived from various sources, including model-based direct and indirect age-specific mortality estimates, given the lack of robust vital statistics systems. This can impact the precision and reliability of mortality estimates despite using sophisticated demographic or statistical modeling to derive them. Second, the period cut-offs were chosen to reflect broad policy and social-epidemiological trends to highlight differences across countries. However, they do not necessarily capture the nuanced changes and disease patterns in all countries, and a country-specific analysis would be necessary to elucidate the paths of change and the factors influencing the dynamics of life expectancy and lifespan variation. Lastly, while this study is descriptive and outlines the complex relationship between age group contributions and associations in life expectancy and lifespan variation, it does not fully dissect the different patterns or the mechanisms driving these relationships. Future research should further investigate how lifespan variation impacts the attainment of sustainable development goals, as well as influence the emerging trends in mortality within the region (Mejía-Guevara et al. 2020).

### Conclusions

Sub-Saharan Africa has experienced a unique pattern of mortality transition unlike that seen in other regions. In the mid-20th century, there were initial improvements in mortality among the young and middle-aged. These were interrupted by significant disruptions during the last two decades of the century, predominantly due to epidemics, especially HIV/AIDS, violence, and social unrest. However, the past twenty years have seen notable improvements, primarily due to the introduction of impactful programs and policies that have accelerated child survival and reduced deaths from HIV/AIDS, malaria, and other infectious diseases. During this transition, improvements in child survival has played a critical role in shaping the current patterns of life expectancy and lifespan variability, while gains at older ages have been slower or uneven. This paper emphasizes the importance of monitoring life expectancy and the variation in age at death to better assess mortality reduction and to set more informed goals for the future.

## Data Availability

All data produced in the present work are contained in the manuscript

