## Supplementary material for "Evolution of Life Expectancy and Lifespan Variation in Sub-Saharan Africa": SupplementaryMaterial_medRxiv_LifeExpectancyTrendsSSA.pdf

### Supplementary Figures

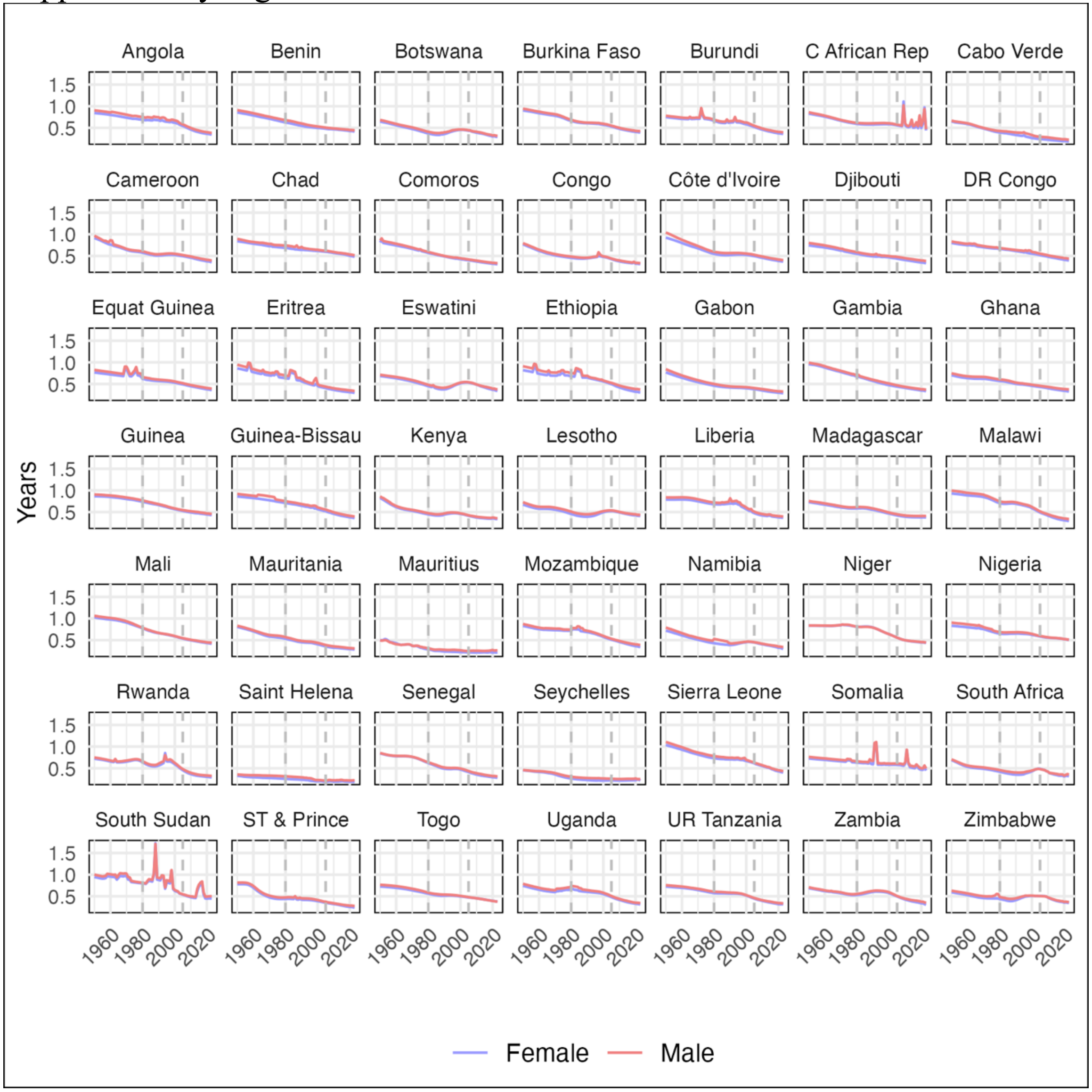


**FIGURE S1.** Coefficient of Variation ($CV_0$) Trends by Gender in 49 Sub-Saharan African Countries, 1950–2023

**Source:** Authors' calculations using data from the UN World Population Prospects 2024 Revision.

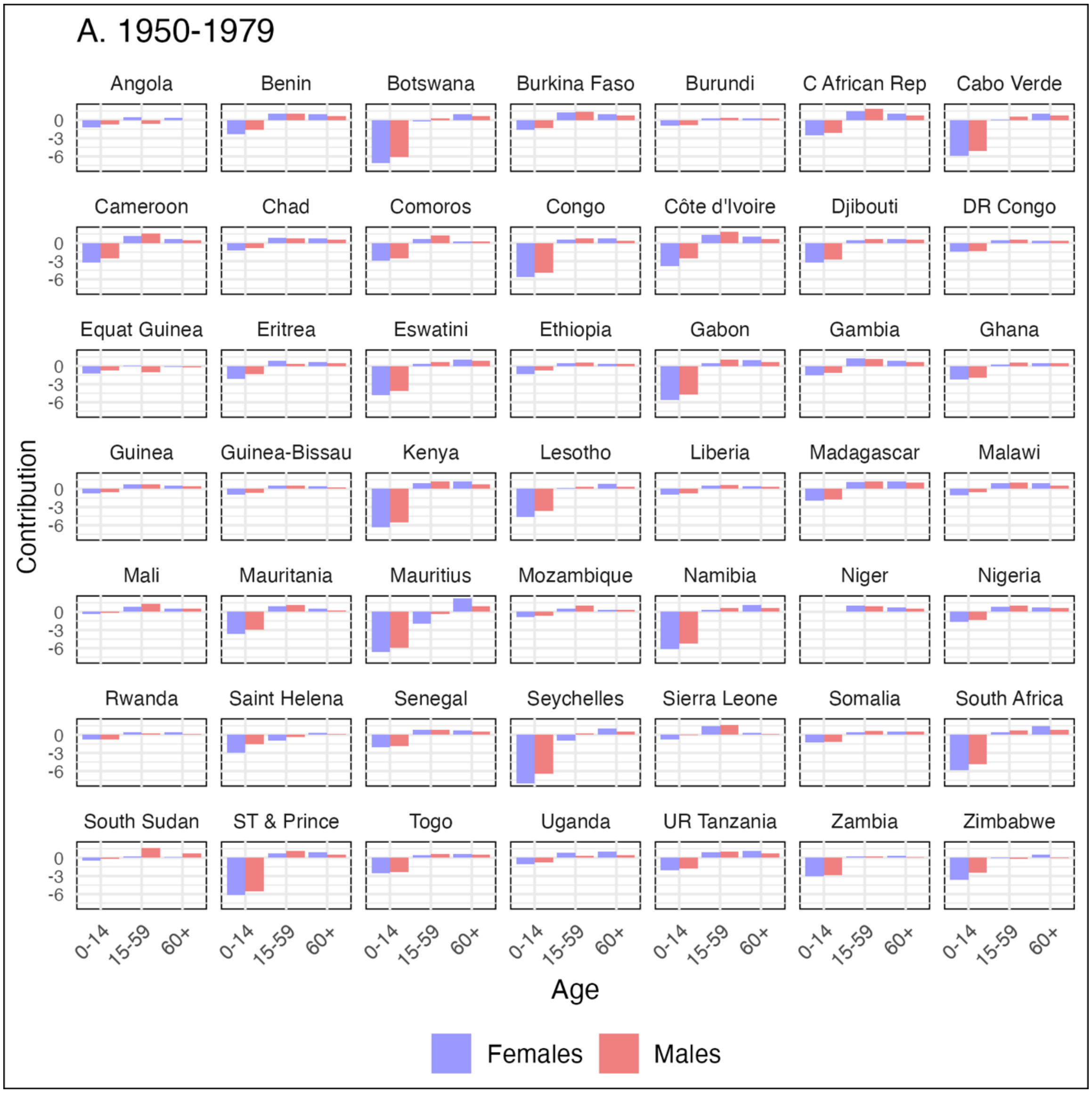
A. 1950-1979
Angola
Benin
Botswana
Burkina Faso
Burundi
C African Rep
Cabo Verde
Cameroon
Chad
Comoros
Congo
Côte d'Ivoire
Djibouti
DR Congo
Equat Guinea
Eritrea
Eswatini
Ethiopia
Gabon
Gambia
Ghana
Guinea
Guinea-Bissau
Kenya
Lesotho
Liberia
Madagascar
Malawi
Mali
Mauritania
Mauritius
Mozambique
Namibia
Niger
Nigeria
Rwanda
Saint Helena
Senegal
Seychelles
Sierra Leone
Somalia
South Africa
South Sudan
ST & Prince
Togo
Uganda
UR Tanzania
Zambia
Zimbabwe
0
-3
-6
Contribution
0-14
15-59
60+
Age
Females
Males

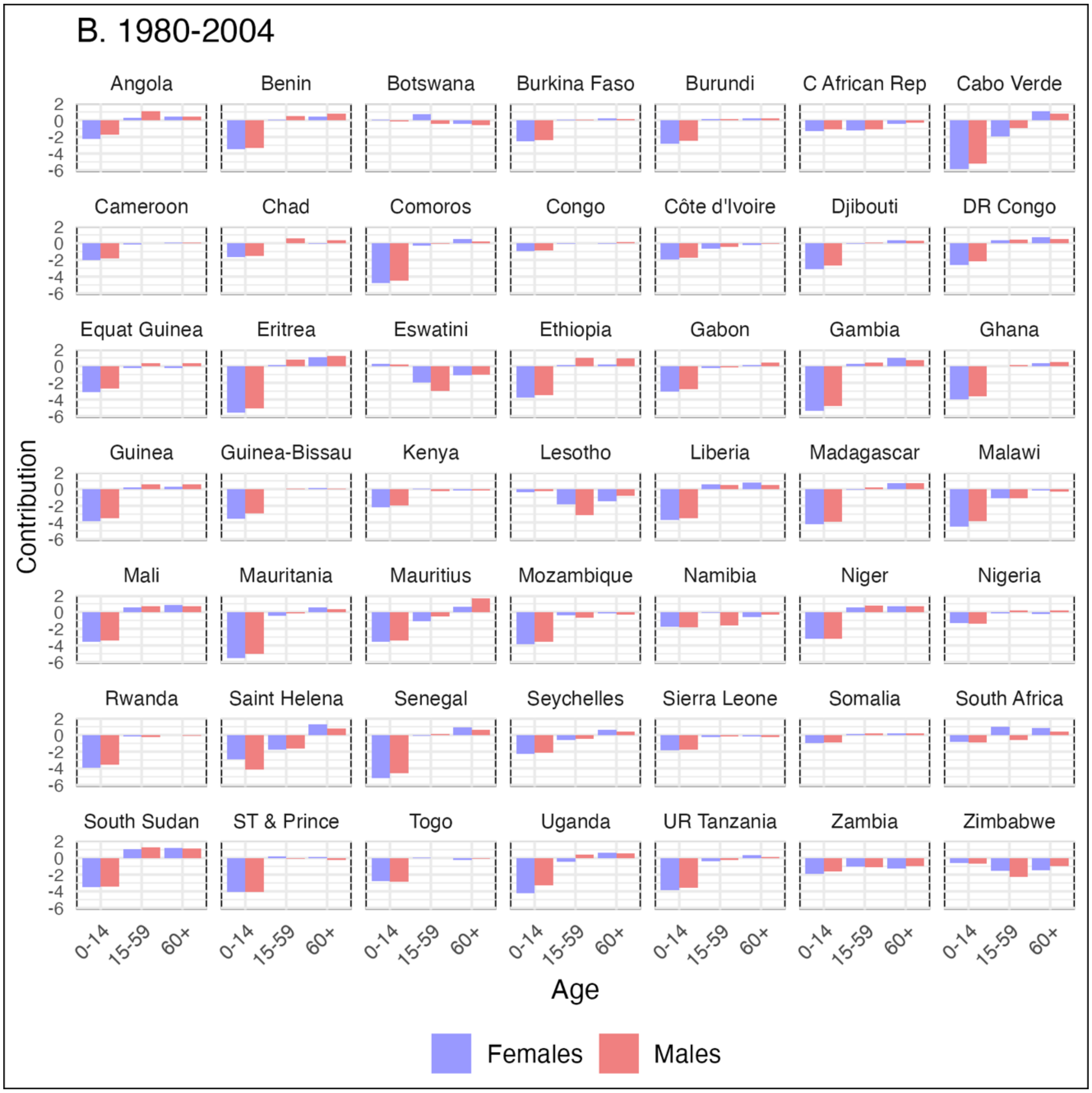

B. 1980-2004
Angola
Benin
Botswana
Burkina Faso
Burundi
C African Rep
Cabo Verde
Cameroon
Chad
Comoros
Congo
Côte d'Ivoire
Djibouti
DR Congo
Equat Guinea
Eritrea
Eswatini
Ethiopia
Gabon
Gambia
Ghana
Guinea
Guinea-Bissau
Kenya
Lesotho
Liberia
Madagascar
Malawi
Mali
Mauritania
Mauritius
Mozambique
Namibia
Niger
Nigeria
Rwanda
Saint Helena
Senegal
Seychelles
Sierra Leone
Somalia
South Africa
South Sudan
ST & Prince
Togo
Uganda
UR Tanzania
Zambia
Zimbabwe
Contribution
2
0
-2
-4
-6
0-14
15-59
60+
Age
Females
Males

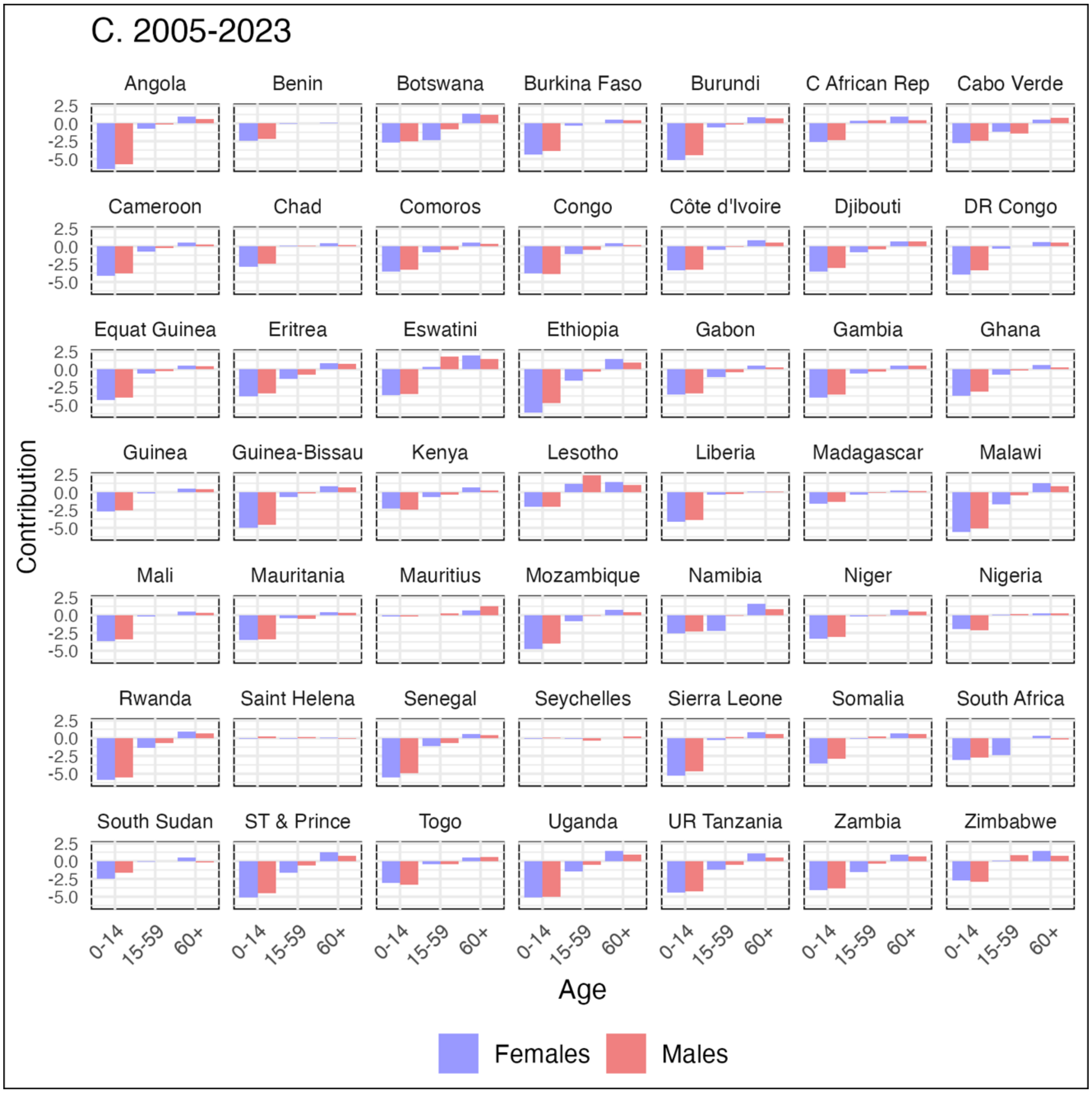


**FIGURE S2.** Age-Specific Mortality Decomposition of Lifespan Variation by Time Period, and Gender in 49 Sub-Saharan African Countries

Panel A: 1950–1979, Panel B: 1980–2004, Panel C: 2005–2023; Age Groups: 0–14, 15–59, 60 and over

**Source:** Authors' calculations using data from the UN World Population Prospects 2024 Revision.
